# The Bambisana Study Protocol: A mixed methods pre- and post- test study assessing community and social media influence to increase influenza vaccination uptake among youth in Soweto, South Africa

**DOI:** 10.1101/2024.03.25.24304871

**Authors:** Janan J. Dietrich, Catherine Hill, Gugulethu Tshabalala, Tshepiso Msibi, Stefanie Vermaak, Nellie Myburgh, Sarah Malycha, Izzy Goldstein, Elliot Grainger, Prima Alam, Kimberley Gutu, Kennedy Otwombe, Heidi J. Larson, Ziyaad Dangor

## Abstract

**Background:** Seasonal influenza has an estimated global reach of 3 to 5 million infections with 290 000 to 650 000 influenza-related deaths yearly. Despite its efficacy in reducing morbidity and mortality, influenza vaccination rates remain low globally and in South Africa. Youth between the ages of 18-34 years are not prioritised for influenza vaccines although influenza surveillance in South Africa shows that individuals aged 19 to 44 present the highest asymptomatic episodes and the lowest medically attended illness. This creates an opportunity to investigate if and how vaccine demand can be created in the absence of clear imperatives to vaccinate. The study tests the effectiveness of tailored, context-specific education, community engagement, including community and social media to increase influenza vaccination uptake. <u>T</u>ailored, context-specific education, community engagement, reliable vaccine supply and free, localised access are all critical for improving perceptions of, increasing confidence in, and motivating uptake of vaccination. This study explores strategies to increase vaccine uptake amongst marginalised youth 18-34 years old in Soweto, South Africa, where influenza vaccines are not universally accessible through the public health system for this age group.

**Methods:** The Bambisana Study uses an innovative approach - including community influencers and social media - to increase uptake of influenza vaccines through designing and testing an integrated communications strategy targeted at marginalised youth in Soweto, South Africa. The Bambisana study uses a mixed methods pre-test, post-test intervention design to test the effects of the interventions.

**Conclusion and Significance:** Enhancing perceptions of, bolstering confidence in, and fostering uptake of vaccination relies heavily on the efficacy of yearly influenza vaccination initiatives, personalized education tailored to specific contexts, active community involvement, consistent vaccine availability, and easily accessible, cost-free distribution channels at the local level.

## Introduction

Globally seasonal influenza causes an estimated 3-5 million infections, and 290 000-650 000 deaths annually [1, 2]. Estimates for influenza burden of disease are limited in Africa; however, hospitalisation and mortality are higher in low- and middle-income countries (LMICs) than in high-income countries (HICs) [3, 4]. In South Africa, an estimated 6 000-11 000 people die from influenza every year, with approximately half of these deaths in the elderly, and people living with HIV [5]. Seasonal outbreaks of influenza pose challenges to the healthcare system, leading to increased hospitalizations, outpatient visits, and strain on healthcare resources [6]. Vulnerable populations, including the elderly, young children, pregnant women, and individuals with underlying medical conditions are at highest risk of severe infection [7].

While non-pharmaceutical interventions can play a role in decreasing influenza infections, vaccination is deemed pragmatic and endorsed by the World Health Organization (WHO) as the recommended method for preventing seasonal influenza [1]. Overall vaccine efficacy to prevent influenza infection ranges from 40-60%, with higher vaccine efficacy (82%) in reducing severe illness and hospitalization [8, 9]. Therefore, the influenza vaccine is especially recommended for those who have chronic illnesses, those who are pregnant, and the elderly [10-12]. Influenza vaccination campaigns typically prioritize these groups during the influenza vaccination season [7, 13]. However, an investigation into the seasonal influenza burden in urban and rural areas in South Africa showed that children younger than 12 years and those 19 and 44 years have an elevated risk of acquiring seasonal influenza [14]. Whilst severe disease is less likely, vaccinating younger adults has the added indirect benefit of providing herd immunity to the family and communities [15, 16]. Furthermore, vaccination offers the potential economic benefit related to the out-of-pocket expenses incurred by patients to treat influenza related symptoms. An estimated annual cost of $111.3 million is incurred by the government, while out of pocket costs incurred by the patient were reported to be $40.7 million [17]. Individuals are at risk of financial loss due to absence at work and missed opportunity for those who are self-employed [18]. Despite recommendations, seasonal influenza vaccine uptake, in the most vulnerable groups, is low in LMICs, including South Africa [19]. Accessibility, lack of knowledge and risk perception have been cited as the most significant barriers to vaccination in South Africa [20].

Despite the demonstrated effectiveness of vaccines in significantly reducing morbidity and mortality of affected individuals, youth are less proactive in health-seeking behaviours due to low risk perception, are not necessarily encouraged to vaccinate for influenza and other infectious diseases [9, 21-23]. For COVID-19, youth were not initially prioritised for vaccination roll-out yet they might have been in contact with vulnerable populations [24, 25]. Additionally, evidence suggests that low vaccination rates are in part due to low health-seeking behaviours among youth. Health-seeking behaviours are the actions involved in achieving and maintaining a healthy condition. [26, 27]. Various factors, both individual and related to healthcare providers, can influence young people’s healthcare-seeking behaviours [28].

Targeted health promotion interventions can increase health-seeking behaviour and uptake of vaccination [29]. Influenza surveillance shows that individuals aged 19 to 44 present the highest asymptomatic episodes and the lowest medically attended illness [14, 30]. Studies undertaken in sub-Saharan Africa have shown that leveraging community involvement in immunization programs positively contributes to vaccine confidence and uptake [31, 32]. In the case of HIV, for example, awareness campaigns to lower the burden of HIV and AIDS epidemics in Southern African countries have proven vital in reducing the rise of disease [33]. The use of television series to promote positive sexual health and prevent HIV effectively engaged the youth by challenging old models of education [34]). The use of social media campaigns have also been increasingly used to promote behaviour change [35, 36], while sparking debate, especially when considering the abundance of false and misinformation [37]. It is vital to consider the significance of social media to improve vaccine uptake in LMICs.

Promoting health care through various platforms and to youth in LMICs may improve uptake of vaccination. Therefore, as part of the Bambisana Project, which includes a diverse and multi-disciplinary team comprising social and behavioural scientists, biostatisticians, population health surveillance specialists, health communications researchers, community influence and engagement practitioners, and marketing and communication strategists, we aim to:

(1) Evaluate the impact of influencer’s online, offline, and in collaboration, on influenza vaccine uptake, with a specific focus on young people, and assess the additional value added by social media engagement;
(2) To design and implement an integrated communications strategy centred around community influencers and social media to increase the uptake of influenza vaccines, particularly among younger populations who are likely unemployed, partially or informally employed and living in Soweto, South Africa;
(3) To explore motivations and barriers to health priorities, and to influenza vaccination, through the use of micro-influencers both online and offline in Soweto, South Africa.

The outcomes of the project will include increased influenza vaccination rates at the local healthcare clinics linked to campaign interventions, alongside an improved attitude towards the importance of vaccination in the target audience - this will be assessed using the Vaccine Confidence Index scale [38]and measured against the most recent Vaccine Confidence Index data from South Africa [39]. Additionally, the project will provide important health systems-strengthening recommendations which could potentially be implemented within current infrastructure.

## Materials and Methods

### Research Design

The primary research aim uses a prospective mixed methods study design including qualitative and quantitative data collection, to assess the effects of community and social media interventions to increase influenza vaccination uptake among youth in Soweto, South Africa. During the intervention phase, surveys will be conducted among clinic attendees accepting as well as declining influenza vaccination within the study clinics.

### Study setting

The study is conducted in Soweto (South-Western Townships) which is a congregation of 29 townships within the Johannesburg Metropolis in South Africa [40]. Soweto is one of the poorest areas of Johannesburg with high rates of youth unemployment which is at 46.3% among the 15-34 year age group [41]

### Sampling

The Bambisana Project leverages the Soweto Health Demographic Surveillance System (HDSS), led by Wits VIDA since 2017 and part of the Child Health and Mortality Prevention Surveillance programme (CHAMPS), provides access to longitudinal health and demographic data for specific enumerated areas of Soweto and the South of Johannesburg, since 2017[42]. The study population includes young adults 18-34 years from four HDSS economically marginalised and geographically distinct Soweto communities: namely Phiri/Senaoane and Mapetla, Mofolo and Meadowlands, Thulani, and neighbouring Thembelihle. Adults (18 years and older), living in these clusters or accessing clinic services in the five study clinics (Senaoane Clinic, Mofolo Community Health Centre, Meadowlands Zone 2 Clinic, Siphumlile Clinic, and Thembelihle Clinic) are deemed eligible for the study. The HDSS communities were included as below (Table 1):

**Table 1:** HDSS cluster summary data.

| Cluster # | Communities | Total Population | Youth 18-34 | # households with youth | Total # of households |
| --- | --- | --- | --- | --- | --- |
| Cluster 1 | Mofolo and Meadowlands | 20986 | 5824 | 3478 | 5912 |
| Cluster 2 | Mapetla & Phiri/Senaoane | 23805 | 6801 | 4037 | 6626 |
| Cluster 3 | Thulani | 23711 | 6848 | 4072 | 6262 |
| Cluster 4 | Thembalihle | 19449 | 5648 | 3831 | 6586 |

**Figure 1:**
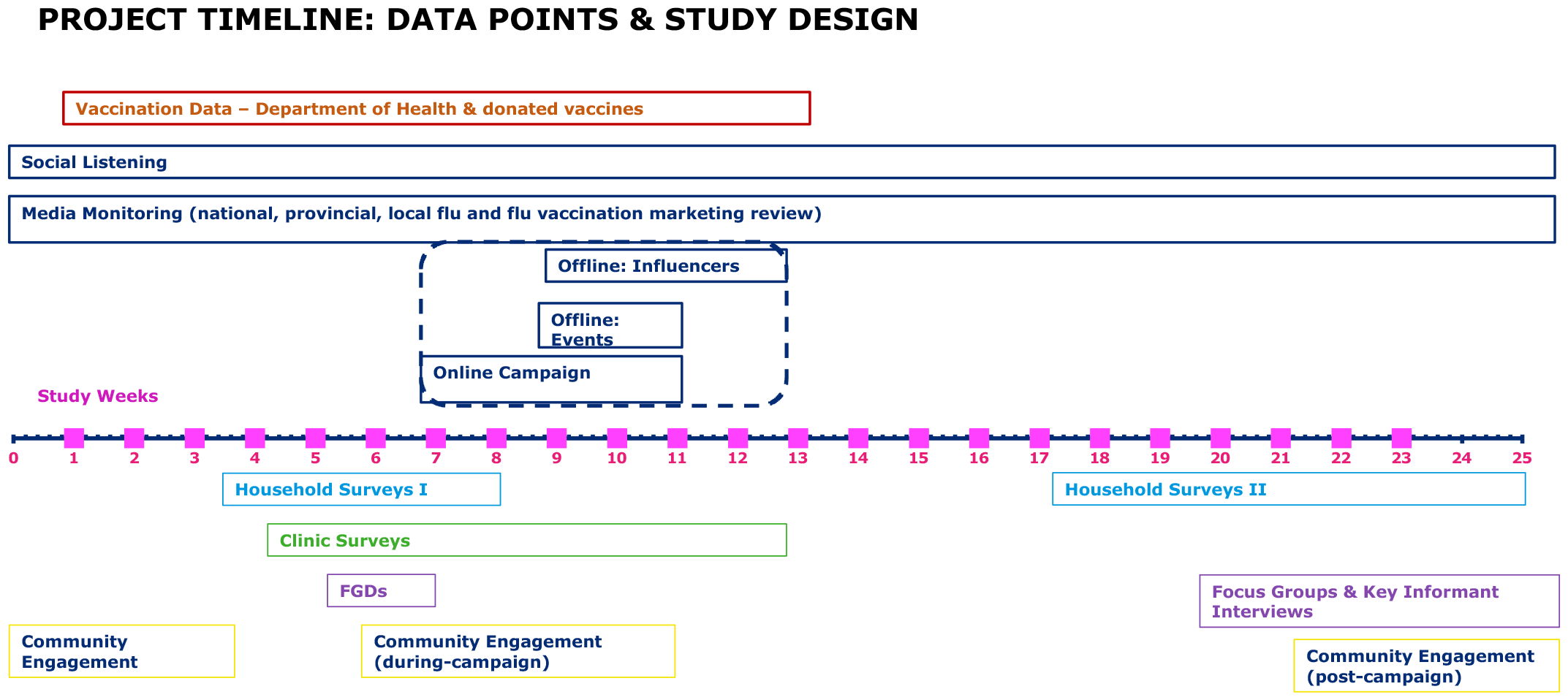
Study Design.

### Community engagement

The Bambisana [an isiZulu word – which means to work together] study leverages active, long-standing, dynamic community-level relationships and partnerships in accessing community insights at the household and local clinic level. The Wits VIDA social behavioural sciences team has longstanding relationships through Community Advisory Boards (CAB) and extensive community engagement in support of health research objectives throughout Soweto. CAB members are individuals residing in the HDSS enumerated areas who have been identified as active citizens in their communities. There are standing quarterly CAB meeting with the study team and members, however, these often increase in relation to shifting project activities. Consultations span project development, implementation, and community feedback sessions at the end of data collection, analysis and interpretation.

### Formative qualitative data collection

The qualitative component includes 16 focus group discussions (FGD) stratified by age group (18-24, 25-34), sex (male and female) and community (two per cluster). These approaches aim to gather contextual data essential for understanding the factors influencing young people’s decisions regarding influenza vaccination, including both motivators and barriers.

Study staff will purposively select and recruit young people for FGDs and key stakeholders/influencers from the community for key informant interviews (KIIs). FGD recruitment will be iterative until data saturation is achieved [43]. All study participants will be recruited through the Community Advisory Board members who work with HDSS communities. Trained and experienced qualitative researchers fluent in local languages spoken in Soweto and Thembelihle (including isiZulu and Sesotho) will conduct the qualitative research using the participants’ language of choice. The FGDs and KIIs will be audio-recorded after written informed consent is obtained.

- FGDs:16 FGDs (8-12 participants per FGD)
- KIIs: 20 (10 male and 10 female participants)

### Intervention

The intervention will be delivered through a public health messaging campaign in targeted clusters or ‘sub-places’ in Soweto, leveraging micro-influencers to provide information and motivate positive attitudes for vaccination without mandate in currently unvaccinated populations. The aim of the campaign is to create a behavioural change action: to go to your local clinic to get vaccinated. This has the secondary aim of highlighting the extent to which such interventions might be possible on wider issues of public health in the future. Secondary to this is the desire to investigate the value-add of social media targeting over wider forms of engagement, such as that which occurs in communities offline.

The campaign will be timed with points when the clinic is well supplied and prepared for such a public drive. The project will mitigate against the risk of potential vaccine shortages by making vaccine donations to partner clinics. The study will centre around the 2023 national influenza vaccination drive in 2023. Vaccines are generally provided in very short and sporadic supply to the local community clinics, so supplying vaccines is also part of the objective to make community-level impact as part of the project.

The intervention will be implemented after community engagement discussions have been conducted to obtain information which would be used to design the messaging and communication strategy. Surveys will be administered prior to, during, and following the communication interventions to establish baseline, midline, and endline measures for evaluating the intervention’s impact. This will help assess the effectiveness of four distinct approaches aimed at boosting influenza vaccine uptake among youth across clinics in four diverse HDSS clusters.

1. **Standard of care:** Vaccine made available with standard government promotional activity without support from the project (i.e., based on standard government drive).
2. **Social media:** Social media campaign to highlight availability and drive to get vaccinated pushed to the target audience.
3. **Micro-influencers on and offline:** The social media campaign will be amplified through a coordinated effort by micro-influencers in their online and offline networks.
4. **Offline micro-influencers**: Non-social media focused; offline micro-influencer led engagement within communities.

The intervention components are detailed below:

#### 1) Social media campaign to highlight availability and drive to get vaccinated pushed to the target audience (under 35s) in four distinct geographic areas

Utilizing Meta platforms Facebook and Instagram, along with other relevant social media channels identified through community engagement and marketing insights, we will disseminate information through static images, carousel posts, and video posts to promote the availability of, education around, and motivation to get vaccinated. Paid advertising on Meta will target people aged 18-34 in two of the four distinct geographic study clusters.

#### 2) Micro-influencers on and offline: The social media campaign will be amplified through a coordinated effort by micro influencers in their online and offline networks

We will use community influencers - people with traditional influence such as community leaders, religious leaders, elders, and key business leaders - to amplify our social media campaign. They will engage their online and offline networks in coordination with our social media campaign to promote vaccine availability and encourage vaccination.

#### 3) Offline micro-influencers: Non-social media focused; offline micro-influencer led engagement within communities

In these communities, our community influencers will focus only on offline engagement and will be specifically instructed to not do anything on social media. Using interpersonal conversations, and small-scale community events to share the importance of vaccines, they will encourage people to get vaccinated.

### Quantitative Data Collection

Quantitative data collection will include five survey data points as outlined below.

All surveys will be developed using the REDCap software, a computer-assisted survey platform with built-in data encryption [44]. All surveys will be completed using dedicated study tablets which means participants would not be required to use their own internet data. Additionally, we will use a data-free survey completion option. The survey links would also be sent to participants who wish to complete the surveys on their own devices. There will be no data costs to participants for completing the assessments. Participants will be reimbursed ZAR 50 airtime towards their time.

A) **Household surveys** include two time points of data collection, a pre- and post-test design, as described below.

### Sampling

Households were chosen through a simple random sampling method, and eligible participants were enrolled as follows: a minimum of 800 adult men and women aged 18 years and above, with 400 designated for the pre-test and 400 for the post-test. At least 100 households with index participants aged 18-34 years will be approached per community. Non-index participants, include other adults in the households who are aged 18-34, 35-49 and 50 years and older who will also be invited to complete surveys.

#### Pre-test and post-test surveys

Using a random sample of households having youth participants (18-34 years) generated from the HDSS database, adults will be approached in their households to complete a 15 minute online pre-test assessment survey to measure: socio-demographics (age, gender, food security, potential loss of income), knowledge of influenza vaccines and benefits, health beliefs and sources of health information (social media, television, radio, friends and family), and vaccine confidence[45]. The same households will be invited to participate in the post-test household survey. The post-test household survey will be conducted after the study campaign intervention.

B) **Clinic surveys** include data collection with vaccinators and those who declined vaccination as described below:

Vaccinated: All adults 18+ years, who received an influenza vaccination will be approached as such numbers may vary as this depends on who elects to vaccinate. However, a total of at least 400 (n=100 per community) youth (aged 18-34) participants who go to the intervention clinics will be purposively approached to complete a post vaccination assessment. The study team will work closely with healthcare workers at public health clinics in the study communities. Healthcare workers will refer patients after having received the influenza vaccine, on site at the clinics. These participants will be invited to complete a brief survey using the same measures as specified for the pre-test survey. The data will be covered as part of the project expenses. There will be no data costs to participants for completing the assessments. Participants will be reimbursed ZAR 50 airtime towards their time.

Additional measures will assess: motivations for influenza vaccines, where participants heard/learned about this or other drives for the influenza vaccination, if participants had seen any online messaging driving them to get the influenza vaccine or been exposed to specific community engagement activities, and whether it had influenced their decision to vaccinate, and online impressions, for example, video views rates (dependent on final campaign strategy and media mix) and their impact on vaccinations in that region. Uptake of influenza vaccines will be tracked in the four clusters across the study period.

**Clinic attendees who decline vaccination** will also be approached to complete surveys. These participants will be 18+ years. Healthcare workers will refer patients who were offered vaccination as part of their clinic attendance but who declined vaccination. Those participants who agree to participate will be invited to complete a brief survey to understand reasons for declining vaccination. The survey measures will include: if and where participants heard/learned about this drive for the influenza vaccination, if participants had seen any online messaging driving them to get the influenza vaccine or been exposed to specific community engagement activities, and whether it had influenced their decision to vaccinate.

As with all the surveys, the data costs will be covered as part of the project expenses. There will be no data costs to participants for completing the assessments. Participants will be reimbursed ZAR 50 airtime towards their time.

C) Qualitative evaluation

Additional FGDs, including 10-12 participants per FGD, will be conducted: 8 mid-way through the intervention (during vaccination season) and 8 post-intervention. These FGDs will be mixed gender and will be stratified by age group (18-24, 25-34), and community (two per cluster). Key informant interviews (KII) (n=20) with key influencers including, religious leaders, traditional healers and youth leaders, as well as other influencers that may identified through community engagement and communications activities.

### Data Analysis

#### Qualitative data preparation and analysis plan

Qualitative data will be analysed using a Framework Analysis approach. Framework thematic analysis provides a highly systematic method of categorizing and organizing data according to key themes, concepts and emergent categories in grids or matrices [46]. The coding will follow a combined approach, i.e. both deductive based on our hypothesized themes and inductive (data-driven) to allow for emerging themes. The analysis will allow us to further refine the domains and will help identify sub-themes. Each transcript will be coded in Dedoose, a qualitative data analysis software by at least two researchers and assessed for consistency in coding of the first 5 transcripts [47]. The research team will convene to review the initial codes and apply general categories that will make up the analytic framework. Any coding disagreements will be discussed until agreement is reached. This analytical framework will then be applied to the remaining transcripts.

#### Statistical Analysis Plan

We will use descriptive statistics to describe study variables including: knowledge of influenza vaccines, experiences of influenza vaccine, stigma and vaccine confidence. Reliability assessments using the Cronbach alpha test will be evaluated for scale measures. Scales with poor reliability scores will be calibrated using factor analysis. Data will be analysed overall and stratified by community cluster. Frequencies will be determined for categorical measures whereas means (standard deviation) and medians (interquartile ranges) will be determined for continuous measures. Pairwise comparisons of categorical measures between communities will be evaluated by Chi-square or Fisher’s exact test as appropriate; Similarly, pairwise continuous measures will be analysed by the two-sample t-test, if satisfying normality distribution conditions or the Wilcoxon Mann-Whitney test, if non-normal. Normality will be assessed by the Shapiro-Wilks method. Drivers of knowledge, stigma and experiences of influenza vaccines will be determined by logistic regression for binary outcomes and linear regression for continuous outcomes. Multivariable analysis will be done to adjust for potential confounders. Model fit statistics will be assessed on all multivariable regression models to ensure the validity of the findings. Data analyses will be conducted using SAS Enterprise Guide 7.15 [48].

### Ethics statement

All research procedures were approved by the University of the Witwatersrand Human Research Ethics Committee (Medical) (M230254) and the local Department of Health. Participants were reimbursed ZAR 50 airtime for the surveys and ZAR 150 for the FGDs and the KIIs.

### Bias correction

While the study is designed to focus on four relatively distinct geographic areas, due to the nature of social media and health-seeking behaviours in the broader community of Soweto, it is possible that members from the intervention communities may interact with those not receiving the intervention. While potential interaction cannot be prevented entirely, research questions for all participants will include questions about exposure to intervention components and other potentially similar national or localised campaigns. In addition, a phased delivery of the interventions to ensure the offline intervention is not biased by the online intervention should help mitigate this.

It is also possible that those exposed to the intervention(s) may seek to be vaccinated in clinics that are not included in the study’s geographically defined target-areas. An effort will be made to collaborate with the Department of Health to broadly understand influenza vaccinations during the campaign season, in broader Soweto and areas where residents typically seek healthcare. Vaccine and vaccination sentiments broadly in Soweto and South Africa will also be tracked during the course of the project for context and monitoring of potential risk.

## Data Availability

Protocol Paper therefore no data reported

## Funding

This project was funded by The Vaccine Confidence Fund II, a philanthropic and charitable fund of Global Impact. Grant ID: VCFII-011. Wits VIDA is an extramural unit of the South African Medical Research Council. The work reported herein for Janan J Dietrich was also made possible through funding by the South African Medical Research Council through its Division of Research Capacity Development under the Early Investigators Programme through funding received from the South African National Treasury. The content and findings reported/illustrated are the sole deduction, view and responsibility of the authors and do not reflect the official position and sentiments of the Vaccine Confidence fund and the SAMRC.

## Acknowledgements

We would like to extend our sincere gratitude to all study participants and beneficiaries, the Wits Vida study team, as well as the FreshNow Team for developing and delivering the content for the campaign.

## Conflicts of Interest

The authors report no conflicts of interest.

## Notes

### Competing Interest Statement

The authors have declared no competing interest.

### Funding Statement

Yes

### Author Declarations

University of the Witwatersrand

